# Multimodal MRI Reveals Brain Structural Differences and Executive Dysfunction in Early Methamphetamine Abstinence

**DOI:** 10.1101/2025.09.15.25335812

**Authors:** Ben Bristow, Maryam Tayebi, Paul Condron, Taylor Emsden, Tutarangi Ngarimu, Eryn Kwon, Rachael McLanachan, Gordon Liu, William Schierding, Patrick McHugh, Samantha Holdsworth, Wendy Mohi, Gil Newburn, Miriam Scadeng

## Abstract

**Background:** Methamphetamine use disorder (MUD) is known to have profound effects on brain structure and cognitive functions. Understanding the extent of these neurobiological changes during the early abstinence period is crucial for developing targeted rehabilitation strategies.

**Objective:** This study aimed to investigate structural brain alterations, and cognitive functions, in early abstinent methamphetamine users compared with healthy controls.

**Methods:** A total of 27 participants were included, comprising 13 participants with MUD in early abstinence (<30 days - mean age: 37.4 ± 9 years, 66% female) and 14 age- and sex-matched healthy controls (mean age: 39.2 ± 11.1 years, 74% female). All subjects underwent brain MRI scans using a 3T scanner, acquiring T1-weighted and myelin-sensitive imaging data. They also completed the Tower of London (TOL) cognitive assessment to evaluate executive functioning. Grey matter volumetric analysis was performed using T1-weighted images, and both regional and global myelin measures were quantified for group comparison.

**Results:** Behaviourally, MUD participants demonstrated significantly longer execution times for correct solutions (p=0.01), required more attempts to solve problems (p=0.02), and achieved fewer correct solutions on the first attempt (p=0.01). Neuroimaging analyses revealed significant cortical thinning in the left lateral occipital and right lingual cortices among MUD participants. Additionally, cortical volumes of the right superior frontal and lingual cortex were significantly reduced in the MUD group. Vertex-wise analysis further showed a negative correlation between duration of methamphetamine use and the volume of multiple cortical regions.

**Conclusions:** These findings indicate that methamphetamine use is linked to poorer executive function, and cortical changes, while the absence of group differences in myelin measures suggests that grey-matter is more vulnerable to meth-induced damage than white-matter.

## Introduction

Methamphetamine is a highly addictive stimulant affecting ~30 million users worldwide (United Nations, 2024) and in 2022 ranked as the third most used drug globally behind cannabis and opioids. As a dopamine agonist, methamphetamine increases synaptic dopamine release and blocks reuptake (Volkow, 2013). It also impacts the broader neurotransmitter systems (Cechova, 2021) by overstimulating postsynaptic neurons and blunting sensitivity to natural rewards, thus reinforcing addiction (May, 2020). Chronic use leads to metabolic, structural, and functional brain changes, contributing to memory deficits, emotional dysregulation, and impaired executive function (Thompson, 2004; Love, 2024). These changes are driven by neurotoxic cascades involving oxidative stress, neuroinflammation, and excitotoxicity, with microglial and astrocytic dysfunction playing key roles (Jayanthi, 2021).

Magnetic resonance imaging (MRI) provides non-invasive insights into brain changes associated with methamphetamine use (Caviness Jr, 1999). Advances in imaging, including high-resolution T1-weighted and myelin-sensitive sequences, now allow detection of subtle alterations in cortical thickness, deep grey matter (GM) volumes, and myelin composition. These advancements allow visualisation of methamphetamine-induced neurotoxicity in regions tied to cognitive dysfunction.

Methamphetamine-related neurotoxicity particularly affects dopamine-rich regions (Ares-Santos, 2013). Using MRI, areas including the striatum (Chang, 2007), prefrontal cortex (Jan, 2012), anterior cingulate cortex (London, 2015), and hippocampus (Golsorkhdan, 2020) have consistently shown alterations. These regions are critical for motor control, executive function, habit formation, reward, and learning (Hardwick, 2013; Diamond, 2013; Graybiel, 2015; Berridge, 2015; Squire, 2015). Particularly, dysfunction in the prefrontal cortex manifests as impulsivity (Kogachi, 2016), poor inhibitory control (Jentsch, 2014), and impaired planning and decision-making (Kohno, 2014), which are common impairments in methamphetamine users. Cognitive impairments, especially in executive control and impulsivity, hinder abstinence (Dawe, 2004). Tests such as the Tower of London (TOL), Stroop task, and Iowa Gambling Tasks have been used to quantify these deficits (Chang, 2002), with TOL performance closely linked to planning ability (van der Plas, 2008).

Beyond GM atrophy, myelin-sensitive imaging techniques such as MAGiC (Nassar, 2023) and diffusion MRI (dMRI) (Aung, 2013) offer insight into myelin and white matter integrity. Myelin facilitates rapid electrical conduction along axons and supports neuronal metabolism (Faria Jr, 2018). Myelin’s involvement in normal neuronal function is well established (Khelfaoui, 2024), reflected by the diverse range of psychiatric and nervous system disorders that are characterised by damaged myelin (Fields, 2008). Methamphetamine use is known to induce chronic neuroinflammation via microglial activation (Sekine, 2008) and oxidative stress responses (Ravera, 2015), both of which may indirectly contribute to myelin damage (Ding, 2021). Disrupted myelin integrity in methamphetamine users is supported by dMRI studies showing increased radial diffusivity, a marker of myelin damage (Liu, 2025; Alicata, 2009). A mouse study by Bowyer et al. (2008) observed myelin damage in the caudate nucleus, and Sharma et al. (2009) identified disrupted myelinated fibres partially in the cortex, hippocampus, thalamus, and hypothalamus through immunostaining in rats both from acute methamphetamine exposure. This supports the findings that methamphetamine exposure can disrupt integrity and potentially damage myelin.

This study investigates whether significant differences in GM volume and myelin content exist between methamphetamine users and healthy controls, alongside assessments of planning and executive cognitive functions. Identifying structural targets of METH neurotoxicity may inform future interventions aimed at supporting grey and white matter recovery during abstinence.

## Methods

### Participants

The participants for this study were recruited through local rehabilitation centres from Tairāwhiti (Gisborne), New Zealand. Thirteen participants who met DSM-V criteria for methamphetamine use disorder (MUD) and had been abstinent for less than one month were included. The control group consisted of 14 participants (age, sex, and ethnicity matched) with no history of methamphetamine use. Ethics approval was obtained (2022 EXP 11360) through the Health and Disability Ethics Committee (HDEC) in New Zealand.

Participants were excluded if they had any contraindications to MRI, had major structural brain abnormalities unrelated to methamphetamine use, a history of other substance use disorders or current intoxication, or any current or prior psychiatric disorders.

### Cognitive test

The TOL task was administered to assess participants’ planning and problem-solving abilities (Shallice, 1982). This involves the movement of balls placed on pegs to match a certain configuration and in a specific number of moves, following the procedure described and discussed in Farhadian et al. (2017, Section 2.3.3) and detailed in our supplementary material. Key metrics are reported in the supplementary material. Additionally, we calculated other parameters including mean first-move time (the average latency before initiating a move, calculated as total first-move time/total attempts), and the impulsivity index (the ratio of mean first-move time to execution time on correct trials).

### MRI acquisition

MRI data were acquired at Mātai Medical Research Institute using a 3.0 T GE Healthcare Signa Premier scanner with a 48-channel Air™ head coil. The imaging protocol incorporated T_1_-weighted BRAVO structural imaging alongside MAGiC, a myelin-sensitive imaging sequence (Table 1).

**Table 1.** MRI acquisition parameters used to acquire 3D T1-weighted and myelin sensitive images.

| Parameter | 3D T1-weighted IR-prep, fast IR-SPGR | 3D MAGiC PROMO |
| --- | --- | --- |
| Repetition Time (TR) | 6.8 ms | 5.72 ms |
| Echo Time (TE) | 2.7 ms | 2.2 ms (max 22.2 ms) |
| Flip Angle | 12° | 4° |
| Matrix Size | 256 × 256 | 256 × 256 |
| Field of View (FOV) | 25.6 cm | 25.6 cm |
| Slices | 320 | 312 |
| Voxel Size | 1 mm<sup>3</sup> | 1 mm<sup>3</sup> |
| Slice Thickness | 0.5 mm | 1 mm |
| Scan Time | 1 min 41 sec | 4 min 49 sec |

### Image processing

All images were visually checked, and those with significant head motion or other artifacts were excluded. The acquired T1-weighted images were processed using FreeSurfer (version 7.4.1, Fischl, 2012) for both cortical and subcortical segmentation.

Myelin-sensitive images were acquired using a MAGiC sequence and processed using SyMRI 15.0 (SyntheticMR, Linköping, Sweden) to generate quantitative myelin maps (Warntjes, 2016). Figure 1 shows an in-depth analysis pipeline overview. The FSL command, *flirt*, was used for linear image registration of the myelin images (Jenkinson, 2001, Jenkinson, 2002).

**Figure 1.**
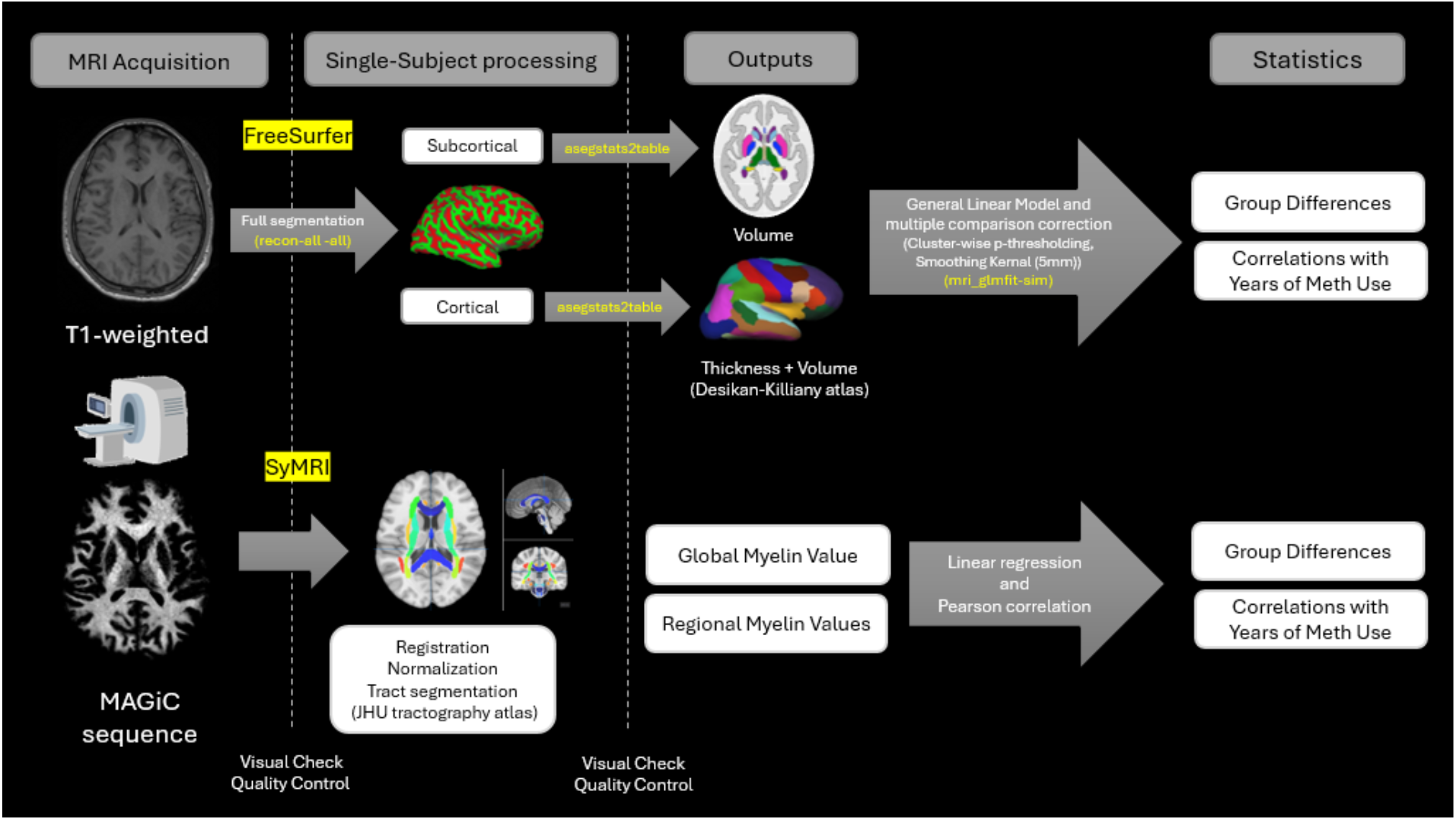
Structural and myelin-sensitive MRI data were processed using two complementary pipelines. T1-weighted images were analyzed using FreeSurfer’s recon-all pipeline to extract cortical thickness, cortical volume, and subcortical volume. The Desikan-Killiany atlas was used for cortical parcellation, and FreeSurfer’s aseg atlas for subcortical segmentation. The -qcache flag enabled smoothed cortical surface data in fsaverage space. Group-level analyses were conducted using mri_glmfit, applying a general linear model with cluster-wise p-thresholding (p < 0.05) and 5 mm smoothing. Myelin-sensitive images were acquired using a MAGiC sequence and processed with SyMRI 15.0 to generate quantitative myelin maps. These maps were registered to MNI152 space using FSL’s flirt, and regional myelin values were extracted using the JHU white matter tractography atlas with fslmaths.

### Statistics

Cortical volume and thickness differences between MUD and control groups were assessed using FreeSurfer’s GLM after surface alignment to a common template and 5mm FWHM smoothing. Vertex-wise analyses included age and sex as covariates, with cluster-wise Monte Carlo correction (p < 0.05). The rest of the statistical analyses were conducted using RStudio (version 2024.12.1+563; R version 4.4.1). Multiple linear regression models investigated group differences in neuroanatomical and cognitive measures - separate models were fitted for each outcome (subcortical volumes, cortical metrics, myelin values, and cognitive indices) - with group (MUD vs. control) as the primary predictor, adjusting for age and sex. For analyses of methamphetamine-use duration, age and sex effects were first regressed out, and the resulting residuals were subjected to Pearson correlations to test associations with brain measures independent of those covariates. All p-values with p < 0.05 considered significant.

## Results

### Participants

The study included a total of 27 participants (13 MUD,14 healthy controls). Shown in Table 2, there was no significant difference in age between the MUD (38.08 ± 9.53 years) and control (39.21 ± 11.13 years) groups. Years of education was significantly higher (t = −3.42, p = 0.0024) in the control group. Participants were predominantly Māori, with a higher proportion of females in both groups; MUD group showed greater rates of head injury, smoking, and poly-drug use, had used methamphetamine for 16.54 ± 21.26 years and had been abstinent for 14.55 ± 5.73 days.

**Table 2.** Participant demographic information.

| Characteristic | Case (N = 13) | Control (N = 14) | p-value |
| --- | --- | --- | --- |
| Age (years) | $38.08 \pm 9.53$ | $39.21 \pm 11.13$ | 0.777 |
| Ethnicity (Maori/NZ European) | 12/1 | 14/0 | 0.481 |
| Sex (M/F) | 5/8 | 4/10 | 0.695 |
| Education (years) | $12.00 \pm 1.73$ | $14.71 \pm 2.40$ | 0.009* |
| Abstinence (days) | $8.64 \pm 9.80$ | | |
| Years of Methamphetamine Use | $14.55 \pm 5.73$ | | |
| Head Injury (Yes) | 5 | 0 | 0.016* |
| Alcohol (Yes) | 6 | 11 | 0.120 |
| Smoking (Yes) | 10 | 5 | 0.077 |
| Poly Drug Use (Yes) | 9 | 6 | 0.322 |

\*= $p < 0.05$

**Table 3.** Cognitive performance on the Tower of London task for MUD and control groups.

| Tower of London performance | Case (M ± SD) | Control (M ± SD) | t-value | p-value | Cohen's d [95% CI] |
| --- | --- | --- | --- | --- | --- |
| Total score | $30.22 \pm 3.53$ | $33.09 \pm 3.70$ | -1.39 | 0.18 | -0.79 [-0.13 -1.71] |
| Mean solution time - correct solutions (ms) | $18770 \pm 6136$ | $22738 \pm 1828$ | -0.40 | 0.69 | 0.28 [-0.61 1.16] |
| Mean execution time - correct solutions (ms) | $8442 \pm 1749$ | $6827 \pm 1100$ | 2.65 | 0.01* | -1.13 [-2.09 -0.18] |
| Total number of attempts | $16.46 \pm 2.66$ | $14 \pm 2.48$ | 2.47 | 0.02* | 0.95 [0.10 1.79] |
| Total first move time (ms) | 164831 ± 56432 | 234779 ± 251643 | -0.94 | 0.36 | -0.40 [-1.21 0.40] |
| Mean first move time (ms) | 10330 ± 4294 | 16661 ± 18324 | -1.17 | 0.26 | -0.50 [-1.31 0.30] |
| Impulsivity index | 0.0019 ± 0.001 | 0.0015 ± 0.0008 | 1.12 | 0.27 | 0.42 [-0.37 1.23] |
| Correct solutions at first attempt | 0.71 ± 0.17 | 0.87 ± 0.16 | -2.5 | 0.01* | -0.96 [-1.80 -0.11] |
| Mean attempts (2-3 moves) | 1.41 ± 0.73 | 1.14 ± 0.36 | 1.24 | 0.22 | 0.45 [-0.35 1.25] |
| Mean attempts (4-5 moves) | 1.47 ± 0.36 | 1.28 ± 0.39 | 1.31 | 0.20 | 0.51 [-0.29 1.32] |

\*=p<0.05

**Table 4.** Correlation results between years of methamphetamine use and cortical volume after adjusting for age and gender.

| Cortical region | Correlation [95% CI] | P-value |
| --- | --- | --- |
| L. Caudal middle frontal | -0.673 [-0.887 0.222] | 0.008 |
| L. Inferior temporal | -0.561[-0.841 0.043] | 0.036 |
| L. Lateral occipital | -0.598[-0.857 -0.099] | 0.023 |
| R. Caudal middle frontal | -0.754[-0.917 0.373] | 0.001 |
| R. Middle temporal | -0.532[-0.829 -0.003] | 0.049 |
| R. Superior parietal | -0.565[-0.843 0.049] | 0.035 |

### Cognitive function

The MUD group showed significantly longer execution times (t = −2.65, p = 0.01), more total number of attempts (t = 2.47, p = 0.02), and less correct solutions at first attempt (t = −2.5, p = 0.01). The TOL performance revealed no significant group differences for total score, mean solution time, total attempts, or first-move time. Although controls scored slightly higher and completed correct trials marginally faster, these differences were minimal.

### Imaging data

#### Group difference - cortical thickness

When comparing cortical thickness group differences, significant clusters were identified in the left lateral occipital cortex (LOC) (peak = 4.287, area = 993.44 mm^2^, cluster-wise corrected p = 0.015) and in the right lingual gyrus (LG) (peak = 4.887, area = 1227.49 mm^2^, cluster-wise corrected p = 0.002), indicating that the control group exhibited significantly greater cortical thickness in these regions compared to the MUD group (see figure 2).

**Figure 2.**
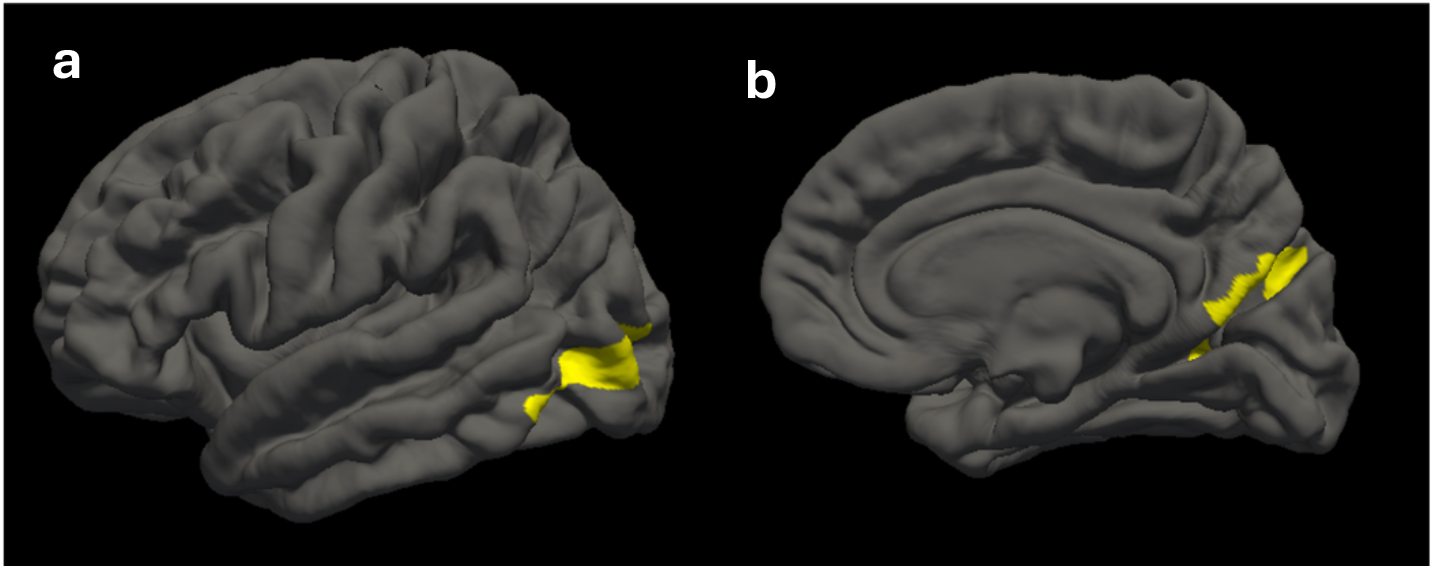
Group differences in cortical thickness between methamphetamine (case) and control participants, identified using FreeSurfer vertex-wise analysis. After regressing out age and sex, the case group exhibited significantly reduced cortical thickness in a) the left lateral occipital cortex (p = 0.015, cluster-wise corrected) and b) right lingual gyrus (p = 0.002, cluster-wise corrected), compared to the control group. The left lateral occipital cortex, involved in object recognition and visuospatial processing, showed reduced thickness in the case group, suggesting possible impairments in visual perception. Similarly, reduced thickness in the right lingual gyrus may reflect disruptions in visual memory and learning associated with methamphetamine use.

#### Group difference - cortical volume

Group comparisons revealed significantly lower cortical volumes in the MUD group than in the control group in two clusters: the right superior frontal cortex (peak = 4.065 within a 1205.87 mm^2^ cluster (cluster-wise corrected p = 0.005)), and the right lingual gyrus (peak = 4.142 within a 1081.3 mm^2^ cluster (cluster-wise corrected p = 0.018)) (see Figure 3).

**Figure 3.**
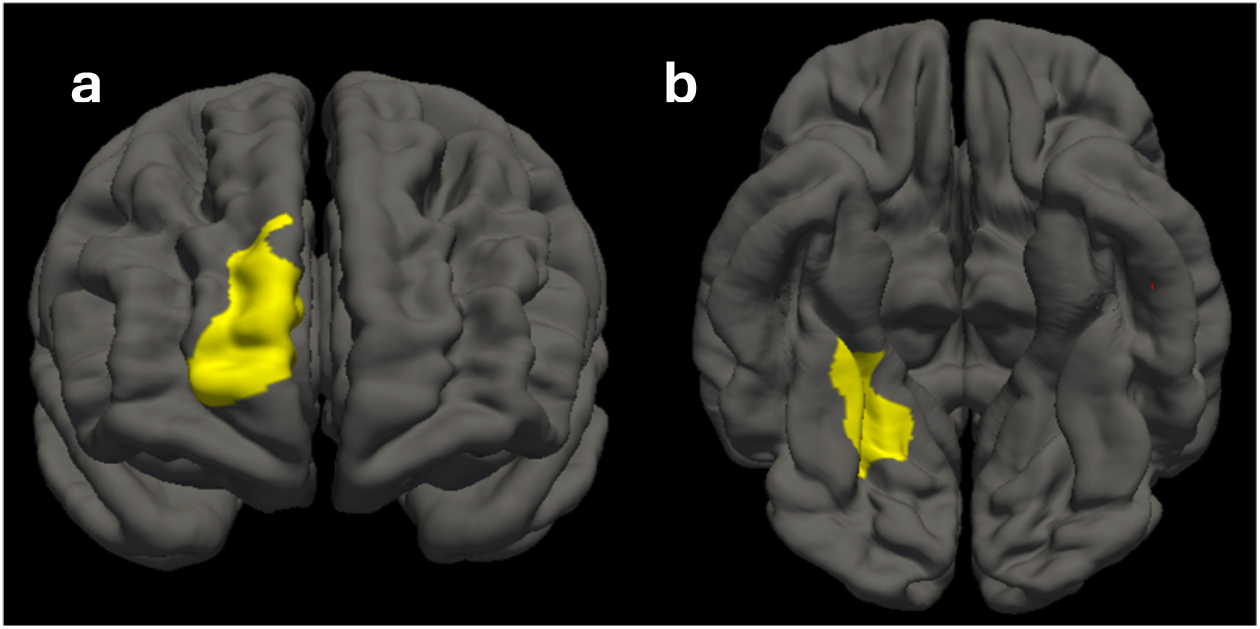
Group differences in cortical volume between methamphetamine (case) and control participants, identified using FreeSurfer vertex-wise analysis. After regressing out age and sex, the case group exhibited significantly reduced cortical volume in (a) the right superior frontal cortex (p = 0.007, cluster-wise corrected) and (b) right lingual gyrus (p = 0.018, cluster-wise corrected), compared to the control group. The right superior frontal cortex, involved in executive functions such as working memory and decision-making, showed reduced volume in the case group, aligning with cognitive deficits linked to substance use. The right lingual gyrus, important for visual processing and memory, also showed reduced volume, suggesting potential impairments in visual information processing among methamphetamine users.

#### Correlation between cortical metrics and years of methamphetamine usage

Cortical thickness did not show any correlation with length of methamphetamine use. Using FreeSurfer’s vertex-wise analysis with cluster-wise correction, we identified a significant cluster in the volume of the left middle temporal cortex that was negatively associated with years of methamphetamine use (peak = –4.40 within a 1,274.10 mm^2^ cluster (cluster-wise correction p = 0.002)). We also performed ROI-based Pearson correlations; as summarised in Table 5, several cortical volumes exhibited moderate, dose-dependent negative correlations with duration of use. The right caudal middle frontal cortex showed the strongest relationship (r = –0.754, p = 0.001), followed by the left caudal middle frontal (r = –0.673, p = 0.008) and the left lateral occipital cortex (r = –0.598, p = 0.023) (Figure 4).

**Table 5.** Myelin content values across various white matter tracts and global myelin volume for the MUD and control groups.

| White matter tracts | Case (M ± SD) | Control (M ± SD) | t-value | p-value | Cohen's d [95% CI] |
| --- | --- | --- | --- | --- | --- |
| Body Corpus Callosum | 23.16 ± 1.95 | 22.82 ± 1.74 | -0.56 | 0.58 | 0.18 [-0.58 0.94] |
| Genu Corpus Callosum | 24.85 ± 2.13 | 24.66 ± 1.89 | -0.24 | 0.81 | 0.09 [-0.67 0.85] |
| Splenium Corpus Callosum | 23.21 ± 1.95 | 23.04 ± 1.95 | -0.27 | 0.79 | 0.09 [-0.67 0.84] |
| L. Superior Longitudinal Fasciculus | 24.57 ± 1.86 | 24.28 ± 1.80 | -0.33 | 0.74 | 0.16 [-0.60 0.92] |
| R. Superior Longitudinal Fasciculus | 23.45 ± 2.00 | 22.98 ± 1.95 | -0.73 | 0.47 | 0.24 [-0.53 1.00] |
| L. Cingulum | 23.83 ± 1.94 | 23.54 ± 1.86 | -0.41 | 0.69 | 0.15 [-0.60 0.92] |
| R. Cingulum | 22.71 ± 2.03 | 22.70 ± 1.94 | -0.08 | 0.93 | 0.01 [-0.75 0.76] |
| L. Superior Corona Radiation | 30.98 ± 2.46 | 30.97 ± 2.49 | -0.21 | 0.84 | 0.00 [-0.76 0.76] |
| R. Superior Corona Radiation | 23.35 ± 1.88 | 22.98 ± 1.85 | -0.66 | 0.52 | 0.20 [-0.57 0.96] |
| L. Anterior Thalamic Radiation | 29.65 ± 2.50 | 30.47 ± 2.45 | 0.69 | 0.50 | -0.33 [-1.10 0.43] |
| R. Anterior Thalamic Radiation | 30.89 ± 2.25 | 30.68 ± 1.88 | -0.12 | 0.90 | 0.10 [-0.66 0.86] |
| L. Inferior Frontal Occipital Fasciculus | 17.46 ± 1.60 | 17.50 ± 1.57 | -0.27 | 0.79 | -0.03 [-0.79 0.73] |
| R. Inferior Frontal Occipital Fasciculus | 31.45 ± 2.19 | 32.10 ± 2.14 | 0.77 | 0.45 | -0.30 [-1.06 0.46] |
| L. Inferior Longitudinal Fasciculus | 31.53 ± 2.56 | 31.91 ± 2.55 | 0.36 | 0.72 | -0.15 [0.91 0.61] |
| R. Inferior Longitudinal Fasciculus | 18.36 ± 1.71 | 18.34 ± 1.63 | -0.16 | 0.87 | 0.02 [-0.74 0.78] |
| L. Uncinate Fasciculus | 22.68 ± 1.82 | 22.52 ± 1.84 | -0.31 | 0.76 | 0.09 [-0.67 0.85] |
| R. Uncinate Fasciculus | 23.45 ± 2.00 | 22.98 ± 1.95 | -0.73 | 0.47 | 0.24 [-0.53 1.00] |
| Global myelin volume | 166.73 ± 22.52 | 172.10 ± 29.87 | 0.23 | 0.81 | -0.21 [-0.99 0.57] |
| Myelin volume (%)<br>(Brain parenchyma normalised) | 13.02 ± 1.47 | 13.15 ± 1.45 | -0.27 | 0.78 | -0.09 [-0.87 0.69] |
| Myelin volume (%)<br>(Brain intracranial normalised) | 11.81 ± 1.53 | 11.87 ± 1.38 | -0.21 | 0.83 | -0.04 [-0.82 0.73] |

**Figure 4.**
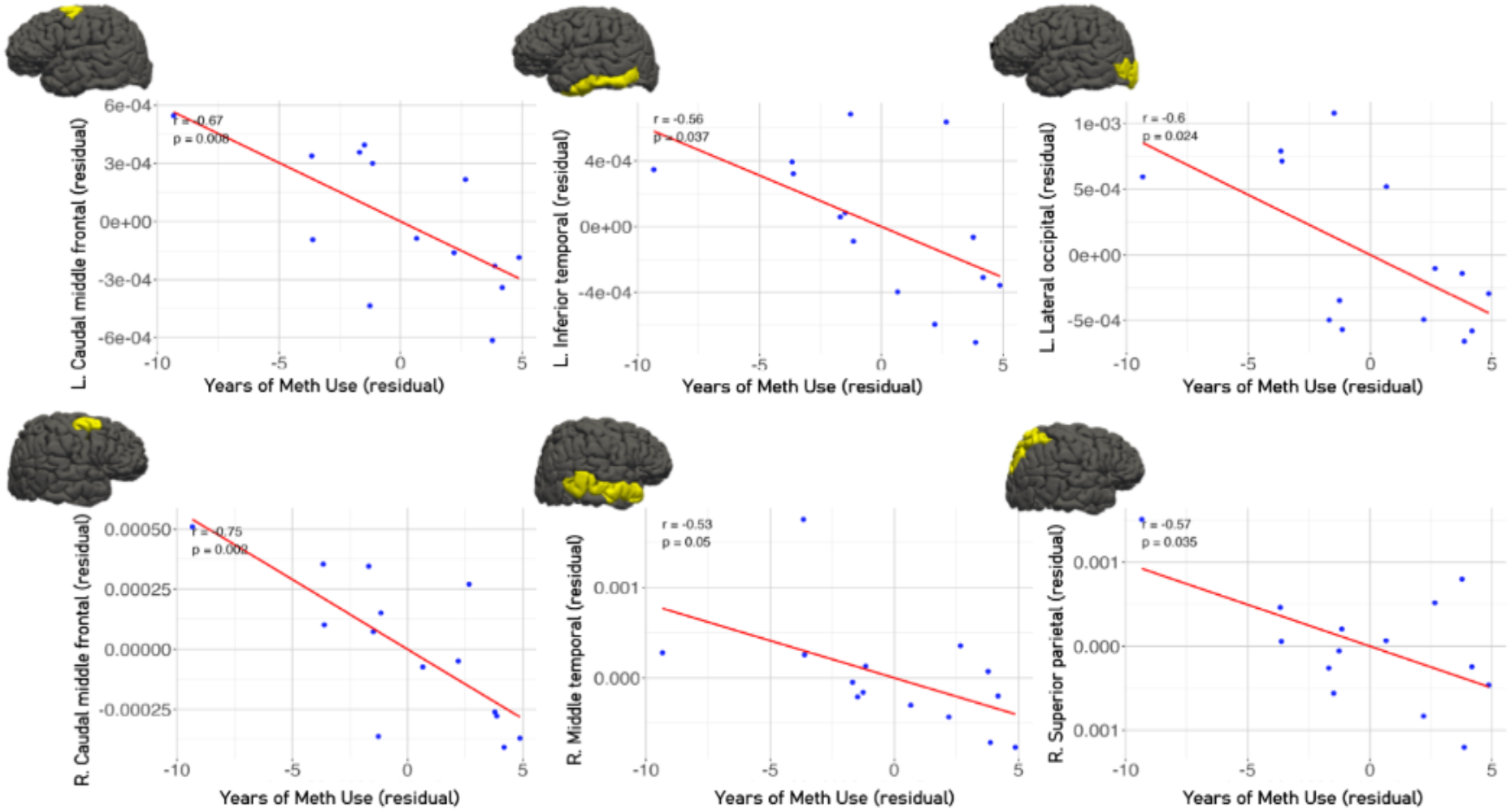
The scatter plots illustrate the negative associations between years of methamphetamine use and cortical volume in specific brain regions, after adjusting for age and gender. These findings indicate that prolonged methamphetamine exposure is associated with reduced cortical volume in regions implicated in executive function and visual processing.

### Myelin

Analysis of myelin content across various white matter tracts revealed no significant differences between the MUD and control groups (Table 6). Similarly, cortical myelin volume did not differ significantly between groups. Although the global myelin volume appeared slightly reduced in the MUD group (M = 166.73 ± 22.52) compared to controls (M = 172.10 ± 29.87), this difference was not statistically significant (p = 0.81) and once corrected for intracranial volume and brain parenchyma this effect diminished. The correlational analysis revealed no significant association between myelin values in any white-matter region and the duration of methamphetamine use.

## Discussion

This study investigated methamphetamine-related brain structural alterations and executive dysfunction, recruiting individuals in early abstinence (<1 month) for multimodal MRI and Tower of London task assessment. MUD group exhibited focal cortical thinning and dose-dependent volume reductions, especially in frontal and occipital regions, along with impairments in executive and other cognitive functions. No significant differences emerged for myelin content or subcortical volume.

### Posterior vision region

Significant thinning was found in the right LG and left LOC. The LOC encodes object form and feeds conscious object recognition and visuospatial attention, whereas the LG contributes to scene processing, visual memory, and the integration of high-resolution visual detail into semantic representations (Velden, 2023; Zhang, 2016). Damage in either region is therefore expected to degrade the fidelity of visual information available for higher-order planning.

A growing body of multimodal work supports the vulnerability of posterior visual-association cortex to psychostimulant exposure and aligns closely with our LG-LOC findings. MRI volumetry in chronic methamphetamine users consistently shows occipital GM loss: Nakama et al. (2011) reported accelerated volume decline in the occipital lobe relative to controls, while Nie et al. (2020) found lower right lateral occipital volume and linked thinner right LG/pericalcarine cortex to longer duration of use in long-term abstinent users. Similarly, we identified negative correlation between the left lateral occipital volume and duration of methamphetamine use. However, there are studies that showed increased GM density in lateral occipital cortex in MUD individuals (Jan, 2012; Morales, 2013). Functional studies show that methamphetamine and other drug exposures heighten activity or alter structure in occipital and lingual regions, suggesting a convergent drug-related impact on visual processing networks (Van Hedger, 2018; Wang, 2012; Makris, 2008).

The occipital cortex is highly glutamatergic and metabolically active, which may render it particularly vulnerable to meth-induced glutamate dysregulation and excitotoxicity. This, in combination with meth-related vascular and neuroinflammatory mechanisms, likely contributes to the vulnerability to structural and functional damage observed in visual–spatial regions (Halpin, 2014; Canedo, 2021; Self, 2012), although direct measures of methamphetamine-evoked glutamate release in visual cortex are limited. Methamphetamine reliably alters glutamate levels within the striatum (Mark, 2004) and prefrontal cortex (Parsegian, 2014; Wu, 2018), while glutamate/glutamine (Glx) in the occipital cortex is known to be modulated during visual processing (Kurcyus, 2018). These mechanisms may underlie the cortical thinning we observed in the LOC and LG, highlighting the occipital cortex as a critical but underrecognized target of methamphetamine neurotoxicity.

### Prefrontal executive region

Cortical analysis identified two closely related dorsolateral prefrontal (DLPFC) regions that are compromised in very-early abstinent methamphetamine users (Kohno, 2014; Zare, 2021). The reduced volume in the right superior frontal gyrus (SFG), together with the dose-dependent association between years of methamphetamine use and smaller volumes in the bilateral caudal middle frontal gyri (cMFG), indicates DLPFC involvement in a progressive impact of methamphetamine use on this region. (Kohno, 2014; Schwartz, 2012). Together these regions constitute the DLPFC system, which underpins working-memory manipulation, set-shifting, impulse control, and strategic planning (Zhang, 2013; Falques, 2014; Hu, 2016; Ye, 2010).

The frontal cortex is a susceptible region for stimulant neurotoxicity, and structural MRI studies exploring methamphetamine use consistently demonstrate prefrontal cortical compromise (Kohno, 2014). Voxel-based morphometry in active users shows GM loss in both medial frontal (Thompson, 2004) and SFG (Jan, 2012). Short-term abstinent individuals (<6 months) still show reduced density in the right middle frontal gyrus compared with those abstinent longer than 6 months (Kim, 2006). Schwartz et al. reported lower density in left middle-frontal cortex, while Kogachi et al (2018) showed sex-specific effects where male users had relatively larger, and female users smaller, frontal volumes - particularly in the medial orbitofrontal and superior frontal cortices - compared with their respective controls. Kogachi et al, 2018 suggested that sex may modulate the effects of methamphetamine on brain morphometry. We have age- and sex-matched the cohorts as well as adding age and sex as covariates in the appropriate models to attempt to control for the effects of age and sex to better power the analysis to see the subtle cortical differences between MUD and control groups. Emerging evidence points to partial prefrontal recovery, with Nie et al (2020) showing that longer abstinence was linked to thicker bilateral SFG - interpreted as reactive gliosis – and Morales et al. (2012) likewise reporting GM increases in the inferior frontal gyri within the first month, suggesting early and continuing cortical restoration. Collectively, these studies represent a dynamic course in which methamphetamine exposure and early abstinence are marked by pronounced dorsolateral and superior-frontal alteration, whereas prolonged drug cessation may permit partial structural rebound.

### Association with length of methamphetamine-exposure

Within the MUD group we observed a clear correlation between duration of use and –brain volume, where longer drug use was associated with smaller cortical volumes in multiple regions after age and sex adjustment. Cortical volume inversely correlated with years of methamphetamine use, particularly in DLPFC, left LOC, inferior temporal cortex, bilateral middle temporal cortex and right superior parietal lobule, consistent with a dose-dependent degradation of executive–visuospatial networks (Ruan, 2017; Nie, 2020). These dose–atrophy relationships appear in both active users and during early abstinence abstinent for only a few weeks and are also echoed by longitudinal non-human primate data (Howell, 2008). Similar trajectories have been observed in MPTP-administered (1-methyl-4-phenyl-1,2,3,6-tetrahydropyridine) primate models, where the neurotoxin MPTP, which selectively targets dopaminergic neurons and causes neuroinflammation and subsequent GM atrophy, paralleling the progressive degeneration seen in chronic methamphetamine exposure (Jeong, 2018).

Dose-dependent prefrontal atrophy accords with earlier morphometric work. Cumulative lifetime dose predicted reduced GM in the right superior frontal and superior temporal cortex, and the right caudate nucleus (Ruan, 2017). A large voxel-based study of recently abstinent individuals likewise found that years of use correlated inversely with GM density of SFG (Schwartz, 2012). Beyond the frontal lobe, prolonged use has been linked to smaller LG and LOC volumes (Nie, 2020) echoing the occipital correlation detected in our study.

### Myelination

No significant differences in myelin content were observed between the MUD and control groups using synthetic MRI, a novel imaging technique applied in this population. While previous studies using diffusion tensor imaging (DTI) have indirectly suggested myelin disruption—particularly via reduced fractional anisotropy in frontal and parietal regions (McKenna, 2016; Liu, 2025), myelination differences were not observed in our population. DTI infers microstructural and myelin/axonal changes from water diffusion patterns, whereas synthetic MRI directly estimates myelin content based on magnetic exchange between surrounding water compartments. Edema-related changes linked to methamphetamine exposure, such as myelin swelling and disrupted tissue structure (Bowyer, 2008; Sharma, 2009), may interfere with accurate myelin quantification. Additionally, the absence of group differences may reflect limited statistical power due to small sample size. Nonetheless, significant GM loss in methamphetamine users suggests broader neurostructural disruption despite preserved myelin metrics.

### Subcortical structures

Although our study did not identify significant differences in subcortical structures between groups, prior research has consistently documented methamphetamine-related alterations in key subcortical regions, particularly within the striatum (Chang, 2005), hippocampus (Golsorkhdan, 2020), and amygdala (Azimzadeh, 2024). For example, Chang et al. 2005 and Jan et al., 2012 reported enlarged striatal volumes in abstinent methamphetamine users, which may reflect a compensatory neuroplastic response to chronic stimulant exposure. Animal studies further support this, with Jedynak et al. (2007) demonstrating methamphetamine-induced structural plasticity in the dorsal striatum, and long-term potentiation observed in the striatum of methamphetamine-exposed rats (Nishioku, 1999; Furlong, 2017). A recent systematic review highlights inconsistencies in reporting the direction of volumetric change in subcortical structures with methamphetamine exposure (Liu, 2025), our cohort did not show significant differences in subcortical regions.

### Cognitive performance

MUD group showed slower, less efficient and accurate performances on the TOL task, solving fewer problems on the first attempt (71% vs 87%), making more total attempts, and taking approximately 24% longer to execute correct solutions, despite comparable total scores. Suggesting participants are taking longer to formulate a plan and when executing the plan, its often inaccurate. These deficits align with structural reductions in dorsolateral prefrontal and occipital regions, which support executive and visuospatial planning.

Performance diverged most clearly on complex tasks: while both groups performed similarly on simpler problems, MUD group made progressively more extra attempts as task difficulty increased, suggesting impaired cognitive flexibility and working memory. This mirrors findings in other stimulant users, where increasing task demands reveal planning inefficiencies (Ersche, 2006). In contrast, some studies report overall lower TOL scores in recently abstinent methamphetamine users (Farhadian, 2017), a difference not observed here.

Despite slower execution, total problem-solving time did not differ significantly, and MUD individuals showed slightly faster (but less accurate) first moves, hinting at impulsive, inefficient planning. Regions in which we observed volumetric reductions, right SFG and bilateral caudal middle frontal gyri, have been repeatedly implicated in planning and executive control by TOL studies and a meta-analysis (Baker, 1996; Nitschke et al., 2017). This anatomical overlap is consistent with the planning inefficiencies measured on the TOL in our cohort. Additionally, thinning in the LOC and LG, which support visuospatial analysis, may impair the mental manipulation of intermediate problem states. These findings raise the possibility that methamphetamine-related neurotoxicity disrupts the integrity of the inferior fronto-occipital fasciculus (IFOF), a key white matter tract linking occipital and prefrontal regions. Prior diffusion studies in methamphetamine users support this hypothesis, showing altered IFOF microstructure (Huang, 2020; Andres, 2016), which may compromise communication between visual input and strategic planning systems.

### Potential confounding factors

Several co-occurring factors may confound the interpretation of neuroimaging and cognitive findings in this study. i) Tobacco smoking, more prevalent among MUD group in our sample, has been associated with reduced GM volume in the prefrontal cortex and dorsal anterior cingulate (Brody et al., 2004). ii)Cannabis use, also more frequent in the MUD group, has been linked to reductions in frontal and temporal GM (Battistella, 2014), potentially exacerbating methamphetamine-related neurotoxicity. Additionally, iii) attention-deficit/hyperactivity disorder (ADHD) is more common among methamphetamine users (Obermeit, 2013), and both ADHD and methamphetamine use disorder is characterised by disruptions to mesolimbic and mesocortical dopamine systems (Eme, 2017). Structurally, thinner right superior frontal cortices have been reported in children with ADHD (Hai, 2022). iv) Trauma can manifest through various forms of abuse, including physical abuse, emotional or psychological abuse, neglect, and exposure to violence (Felitti, 1998). Particularly in childhood, the neurobiological manifestations of abuse are found to translate into abnormal development of important stress, emotional regulation, cognitive, and reward pathways (Wiet, 2017). These neurobiological changes predispose individuals who have suffered trauma to substance use disorders later in life, including methamphetamine addiction (Banducci, 2014; Rasche, 2016; Miela, 2018). Together, these factors may independently or synergistically contribute to the observed brain alterations, complicating attribution solely to methamphetamine exposure.

## Limitations

The primary limitation of this study is the small sample size, which reduces statistical power and limits the ability to detect subtle or region-specific group differences. This also restricts generalisability. Although group demographics were matched, high rates of polysubstance use, head injury, and psychiatric conditions in the MUD group were self-reported and not clinically verified, introducing potential misclassification bias. The use of self-report measures may result in underreporting or inaccuracies in substance use history and comorbid diagnoses. Finally, while the study controlled for age and sex, other variables such as sleep, diet, childhood trauma, comorbidity, education, and socioeconomic status were not accounted for and may influence brain structure or cognition. These factors should be considered in future studies with larger, more diverse cohorts and standardized clinical assessments.

## Conclusion

Our early-abstinent methamphetamine group showed evidence of impaired planning and GM loss in the dorsolateral prefrontal and occipital cortices, with the severity of structural decline scaling with cumulative drug exposure. This pattern supports a dose-dependent breakdown of fronto-occipital networks critical for executive control. Given the lack of observed myelin and subcortical volume differences, cortical GM may exhibit greater susceptibility to methamphetamine-induced structural alterations than white matter. Future multimodal studies with larger, better-controlled samples are needed to disentangle drug-specific effects from those of polysubstance use, comorbid conditions, and individual neurodevelopmental risk factors.

## Data Availability

All data produced in the present study are available upon reasonable request to the authors

## Acknowledgements

This research was supported by funding from the Fred Lewis Enterprise Foundation and Hugh Green Foundation. The authors gratefully acknowledge their contribution, which made this work possible. WS was supported by a senior research fellowship from the Vision Research Foundation. MT and EK were supported by a senior research fellowship from the Hugh and Moira Green Foundation.

