## Supplementary Material for "Multimodal MRI Reveals Brain Structural Differences and Executive Dysfunction in Early Methamphetamine Abstinence"

**Cognitive test**

Participants were required to rearrange a set of coloured beads across pegs to match a target configuration. Prior to the experimental trials, participants received standardised instructions and completed one practice problem to ensure familiarity with the task demands. The puzzles were presented in a controlled, distraction-free environment.

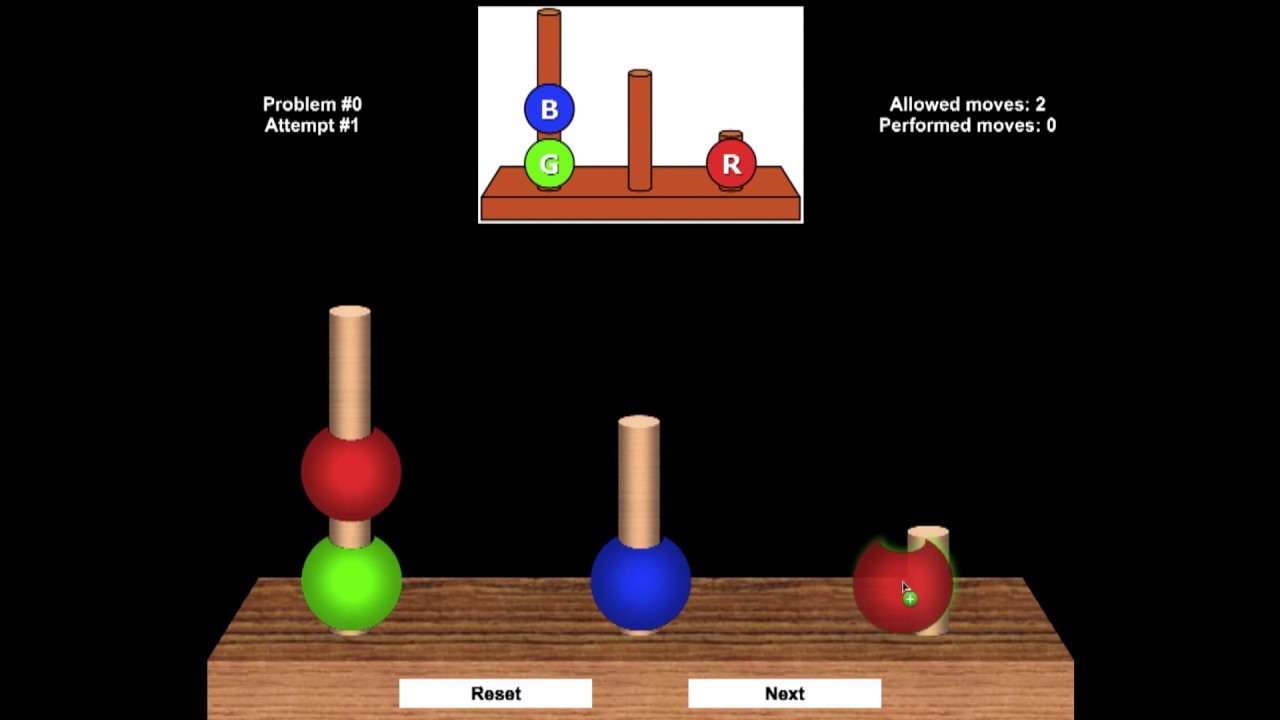

**Apparatus and Stimuli**

The Tower of London task was administered using a computerized interface displaying three vertical pegs of heights 3, 4, and 5 units, and three coloured beads (red, green, blue). Participants manipulated the beads by clicking and dragging with a mouse (or touchpad). All stimuli and responses were presented and recorded using Inquisit 6 Lab (Millisecond Software, Seattle, WA, https://www.millisecond.com/).

**Trial Structure**

A total of 12 planning problems were presented, each defined by a minimum move count between two and five moves. Problems were divided equally into low (2–3 moves) and high difficulty (4–5 moves) and were presented in the same fixed order for each participant. Each trial began with the target configuration displayed above the starting configuration. Participants were instructed to plan ahead and then initiate moves by clicking on the bead to move and the destination peg. If the participant did not complete the problem in 3 attempts, they were moved onto the next problem.

**Practice and Instruction**

Prior to experimental trials, participants received a verbal explanation, standardized onscreen instructions and completed one practice problem (3-move difficulty). Feedback was provided during practice to ensure task understanding; no feedback was given during the main trials.

**Timing and Response Recording**

For each problem, the software recorded:

- Total Score: total score of whole trials
- First-move time: latency (ms) from problem onset to the first click (planning latency)
- Execution time: time (ms) from first click to placement of the final bead correctly
- Solution time: time (ms) to complete a problem correctly
- Total attempts: number of attempts made
- Excess moves: total moves minus minimum moves required

**Derived Metrics**
In addition to the above, we calculated:

- Mean first-move time (total first-move time ÷ number of trials)
- Impulsivity index (mean first-move time ÷ mean execution time on correct trials)
- Mean attempts (2-3 moves)
- Mean attempts (4-5 moves)
